# Sex and ethnicity differences in coronary heart disease: A UK-based tri-ethnic cohort analysis

**DOI:** 10.64898/2026.07.17.26357977

**Authors:** Demelza Smeeth, Sophie V. Eastwood, Andrew Wong, Alun D. Hughes, Nish Chaturvedi

**Author notes:** Corresponding author: Name: Demelza Smeeth, Address: Unit for Lifelong Health and Ageing, UCL, Floor 5, 1-19 Torrington Place London, WC1E 7HB.

## Abstract

**Background and aims:** Sex differences in ethnic minority risk of coronary heart disease (CHD) are often overlooked. Here we aim to explore sex-by-ethnicity differences in CHD outcomes and the contribution of risk factors.

**Methods:** Incident CHD events were identified for Europeans, and South Asian and African/African Caribbean first generation migrants in the UK-based Southall and Brent Revisited (SABRE) cohort. Cardiovascular risk factors were assessed at baseline (1988-91). Cox proportional hazards models quantified group differences in CHD incidence and risk factor contribution. Population attributable fractions (PAFs) described risk factor contribution to group differences.

**Results:** Among 4,754 participants followed for 40.8 years, 1,710 CHD first events occurred. Cumulative incidence of CHD was highest in South Asian males (65% by age 90) and females (55%), compared with 52% in European males and 24-31% in other groups. Sex differences in CHD incidence were pronounced in Europeans (female versus male HR=0.45, 95% CI [0.37,0.55]) but attenuated in South Asians (0.68 [0.56,0.82]) and African/African Caribbeans (0.79 [0.57,1.10]). CHD risk was higher in South Asian compared to European men (1.80 [1.63,1.99]). This ethnic difference was greater in females (2.44 [1.88,3.17]). PAFs for diabetes (PAF=18.0%, 95% CI [6.2,29.8]), hypercholesterolemia (44.2% [20.4,68.1]), and hypertriglyceridemia (22.4% [7.9,37.0]) made a greater contribution to the risk of CHD in South Asian females compared to all other groups.

**Conclusions:** Ethnic minority female participants do not have the same protection from CHD as Europeans. Greater cardiometabolic burden may drive this elevated CHD risk and loss of female protection.

## Introduction

There are marked between ethnic group differences in coronary heart disease (CHD) risk, even within the same geographical setting. In the UK, people of South Asian descent (i.e. with familial origins from India, Pakistan and Bangladesh) experience CHD risks that are 1.5-2 fold that of the general population, while people of African or African-Caribbean ancestry have rates of CHD that are substantially lower than the general population.^1–3^

Sex differences in CHD risk by ethnicity have largely been overlooked.^4^ In people of majority European ancestry, women have about half men’s risk of CHD;^5^ however, whether such sex differences are equally experienced across different ethnic groups is not clear. Similarly, ethnic differences in CHD tend to be examined in combined samples of men and women, ignoring the potential for sex-specific patterns of CHD risk across ethnicities. Limited evidence suggests that sex-by-ethnicity differences may occur for CHD but research is limited to US populations and their main ethnic groups. Findings from the US on ethnicity and CHD risk are also limited in generalisability by inequitable access to healthcare. There is also evidence for sex-by-ethnicity differences in specifically incident acute myocardial infarction (MI) in the Netherlands,^9^ Norway,^10^ Canada,^11^ and Sweden.^12^ However, research is dated, focuses on ethnic groups with limited relevance to other regions of Western Europe, and the use of registry data limits the exploration of the complex risk-factor profiles and contextual influences required to understand sex-by-ethnicity differences in CHD incidence.

In addition to disease risk, common risk factors and their relative contribution to disease incidence may not be experienced equally across groups. Both sex and ethnicity are accompanied by a plethora of social, environmental, biological and genetic differences that may feasibly mediate inequalities in CHD outcomes.^5,13^ Indeed, research indicates that clinically-relevant composite scores of common cardiovascular risk factors may be less relevant for female and non-White populations.^14^ Research that investigates the sex and ethnicity differences in individual risk factor contribution is scarce, but suggests that such group differences may exist for some cardiovascular risk factors and outcomes. An expansion of the risk factors and cardiovascular outcomes studied is warranted.^15,16^

What remains unclear is how the risk for CHD differs by sex and ethnicity across the major UK ethnic minority groups (South Asian, and African and African Caribbean) compared to those of the majority ethnic group (White European). We therefore sought to determine whether a) known ethnic differences in CHD, MI risk, or CHD mortality are equally experienced by males and females, b) whether known sex differences in CHD, MI risk, or CHD mortality in European origin populations would be recapitulated in minority ethnic groups, and c) how common risk factors contribute to CHD across intersectional sex-ethnicity groups.

## Methods

### Cohort

The SABRE study is a tri-ethnic community-based cohort from North-West London. Details of the cohort have been published.^2,17^ Briefly, 4,857 participants aged 40 to 69 years at baseline (1988 through 1991) were selected randomly from primary care physician lists and workplaces in the London districts of Southall and Brent. Ethnicity was agreed on with the interviewer at baseline based on self-report and parental place of origin (European, South Asian, or African and African Caribbean). All South Asian, and African/African Caribbean participants were first-generation migrants. South Asians originated from the Indian subcontinent (the majority, 93%, were from India and Pakistan) and 64% were of Punjabi Sikh descent. African-Caribbeans originated from the Caribbean (91.5%) or from West Africa. Due to small group sizes, we were unable to use more granular ethnic descriptions in analyses. Sex was self-assigned. All participants gave written informed consent. Ethical approval for the study at baseline was obtained from Ealing, Hounslow, and Spelthorne, and University College London research ethics committees.

Baseline assessment comprised in-depth demographic, health and lifestyle questionnaires as well as a clinic visit which collected blood pressure measurements, electrocardiography, anthropometry, fasting blood and overnight urine samples. Since baseline, participants have been flagged for death by the Office for National Statistics and Hospital Episode Statistics were obtained for traced participants.

### Measures

#### Incident CHD events

Our primary outcome of interest was a composite indicator of CHD identified from a combination of primary care, hospital and mortality records. This broad approach was chosen due to the differences in symptom presentation and diagnosis that occur across groups.^18^ We identified the first CHD event in the time since baseline from a combination of follow-up data sources in a similar manner to previous studies.^2^ The first CHD event was identified from: 1) cause of death including myocardial infarction or its sequelae, or atherosclerotic heart disease; 2) primary care record review by two senior physicians according to pre-determined criteria based on symptoms, cardiac enzymes, electrocardiography findings, and hospital discharge diagnosis,^19^ 3) Hospital Episode Statistics mentioning similar conditions to 1, and interventions or rehabilitation for CHD (e.g., angioplasty or coronary artery bypass graft). In addition, we identified acute myocardial infarction events from: 1) underlying cause of death including myocardial infarction; 2) myocardial infarction identified from primary care record review, 3) Hospital Episode Statistics mentioning myocardial infarction, and CHD-related deaths from cause of death including myocardial infarction or its sequelae. Full definitions are available in the Supplementary Methods. The censoring date for all event types was 13^th^ August 2024.

#### Cardiovascular risk factors

A series of cardiovascular risk factors were assessed at baseline (1988-1991). Smoking status was self-reported. Hypertension was identified from self-reported doctor diagnosis or blood pressure (BP) measurements (systolic BP≥140 mmHg or diastolic BP≥90mmHg). Diabetes was identified according to World Health Organization criteria,^20^ self-report of doctor-diagnosed diabetes, or receipt of anti-diabetes medications. This was not limited to type 2 diabetes but is expected to primarily comprise these cases due to general disease prevalence, particularly in this older population.^21^ Anthropometric measures provided measures of central adiposity (waist-hip ratio) and obesity (body mass index; BMI). Central adiposity was defined as a waist-hip ratio ≥0.85 in women and ≥0.90 in men. Obesity was defined as a BMI≥30kg/m^2^. Fasting total cholesterol and triglycerides, were assessed from collected morning blood draws. Elevated total cholesterol (hypercholesterolaemia) was defined as ≥5mmol/L,^22^ while elevated fasting triglycerides (hypertriglyceridemia) were defined as ≥1.7mmol/L.^23^ Overnight urine collections were sampled to capture albuminuria (combining microalbuminuria and macroalbuminuria) representing poor kidney function, which was defined as albumin urinary excretion rate ≥20 μg/min.^24^ Frequency of fruit and green vegetable consumption in the past week was assessed by a simple dietary questionnaire (not in last 7 days, 1 day/week, 2-3 days/week or most days). Low consumption of fresh fruits and vegetables was defined as those who reported eating fresh fruits or vegetables less frequently than most days. Physical activity was calculated as the total weekly energy expended (in megajoules) across sports, walking, and cycling using questions and energy expenditure estimates as described previously.^2^ Area-based deprivation was derived from residential postcode using the Townsend deprivation index.^25^ This was used as a binary variable comparing the most deprived quintile (5) to quintile 1-4.

### Statistical analyses

All analyses were performed using R (v. 4.4) running in RStudio. Analyses were performed in the sub-population with follow-up data who had not experienced a CHD event at baseline (n=4,726). Participants with missing data were excluded from relevant analyses (listwise deletion). All analyses adjusted for age at baseline.

We estimated the cumulative incidence of CHD events, MI events or CHD-related deaths using the cumulative incidence function from the *tidycmprsk* R package.^26^ Cox proportional hazards models were used to evaluate sex and ethnicity differences in incidence of CHD, MI or CHD-related deaths using the *survival* R package.^27^ We also examined a) ethnic differences stratified by sex, and b) sex differences stratified by ethnicity. A likelihood ratio test was used to compare models with and without the interaction term between sex and ethnicity.

We explored the relationship between common CVD risk factors and CHD incidence across each of the six sex-ethnicity strata using Cox proportional hazards models. Risk factors were modelled in separate models adjusting for age at baseline. Risk factors included clinical indicators (diabetes, hypertension), biochemical indicators (albuminuria, hypercholesterolemia, hypertriglyceridemia), biometrics (waist-to-hip ratio, obesity), health-related behaviours (ever smoking, low fruit and veg consumption, low physical activity) and socioeconomic status (area deprivation). All risk factors were used as binary variables to allow for modelling of population attributable fractions (see below).

We used Cox proportional hazards models rather than Fine-Gray competing-risk models because our aim was to estimate etiologic differences by sex and ethnicity, and Fine-Gray sub-distribution hazards can be influenced by group differences in competing mortality, complicating interpretation of the associations with the event of interest.^28^ However, sensitivity analyses were conducted using and Fine-Gray sub-distribution hazards models. Proportional hazards assumptions were assessed for all covariates using Schoenfeld residuals via the *survival* package. Where violations were identified, the nature of the time-varying effect was examined by plotting scaled Schoenfeld residuals against follow-up time using the *survminer* package.^29^ Where the violation was by age at baseline, we replicated models with an interaction between age at baseline and logged time to ensure results were robust. Where the violation was by the covariate of interest, follow-up time was split into intervals (0-10, 10-20, 20+ years) and separate Cox proportional hazards models were fit within each interval, adjusted for age.

Population attributable fractions (PAFs) were calculated to examine the relative contribution of common CVD risk factors to the development of CHD across sex-ethnicity groups. These reflect the relative prevalence of the risk factor as well as the strength of its association with the outcome. PAFs were calculated per strata using *graphPAF* using Cox proportional hazards model-derived hazard ratios, and the direct calculation method across follow-up times of 5, 10, 20, and 30 years.^30^ Although some deviations from proportional hazards were detected in the underlying Cox models, PAFs were highly consistent across follow-up times, indicating that these deviations did not materially influence the estimated attributable risk. Therefore, illustrative PAFs are reported for 20 years of follow-up. Confidence intervals were estimated using bootstrapping up to 500 repetitions. Primary models were only adjusted for age at baseline in order to directly compare a mixture of behavioural, environmental and clinical risk factors, therefore may be confounded and should not be considered to represent the causal contribution of the risk factor to CHD. Follow-up models additionally adjusted for 1) area deprivation; 2) area deprivation, smoking, fruit and vegetable consumption, and physical activity; 3) area deprivation, smoking, fruit and vegetable consumption, physical activity, and obesity.

## Results

### Cohort description

At baseline the cohort comprised 4,857 participants, 26 (0.5%) of whom had experienced a prior CHD event and 4,754 (97.9%) who had at least one form of follow-up data (Supplementary Figure S1). After excluding those with pre-baseline CHD or missing follow-up data, the main analytical sample comprised 4,728 participants: 1,751 European males, 1,377 South Asian males, 439 African/African Caribbean males, 551 European females, 286 South Asian female, and 324 African/African Caribbean female (Table 1). The mean age of participants was 52.2 years (SD: 6.9).

**Table 1.**
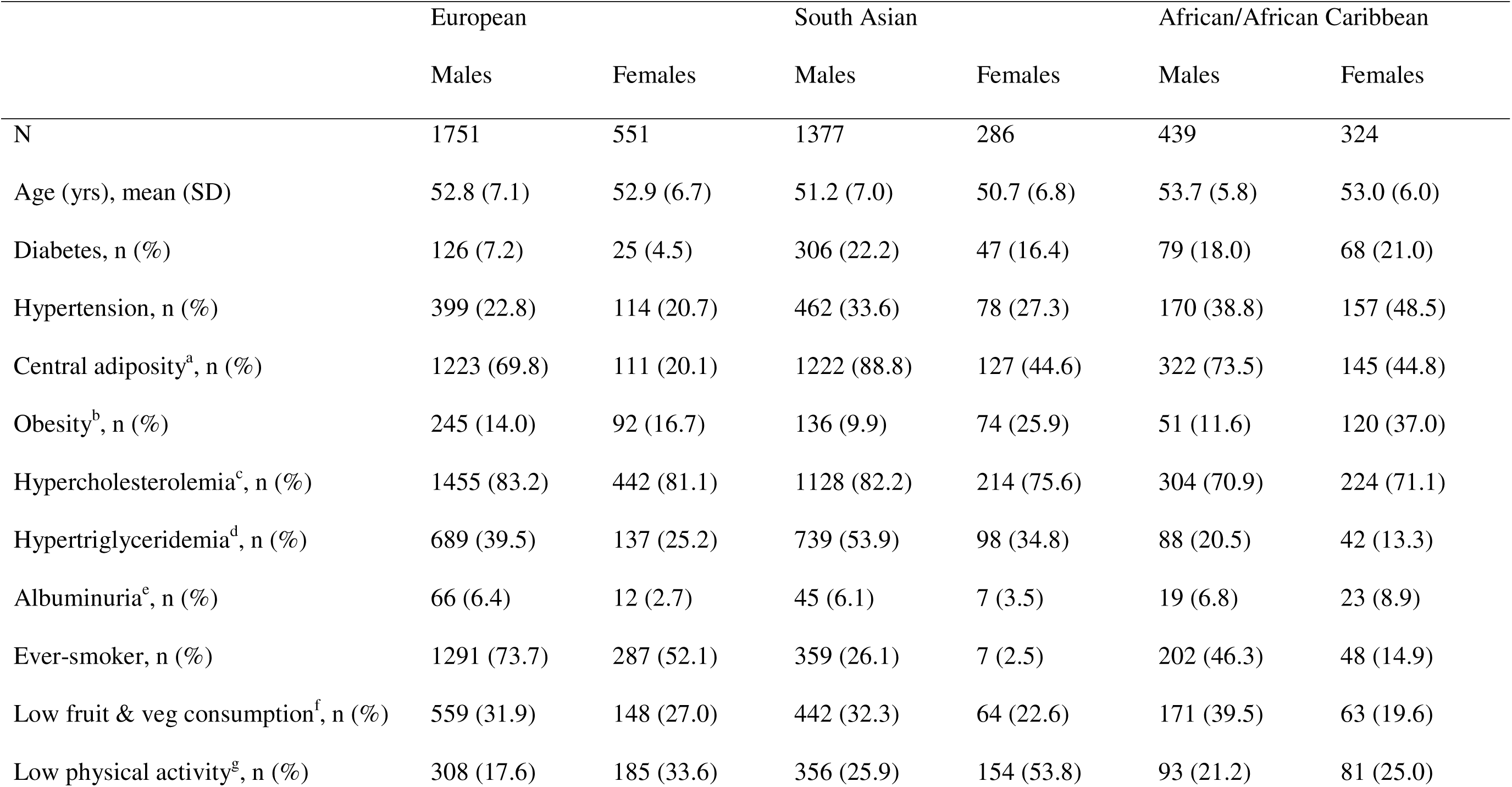
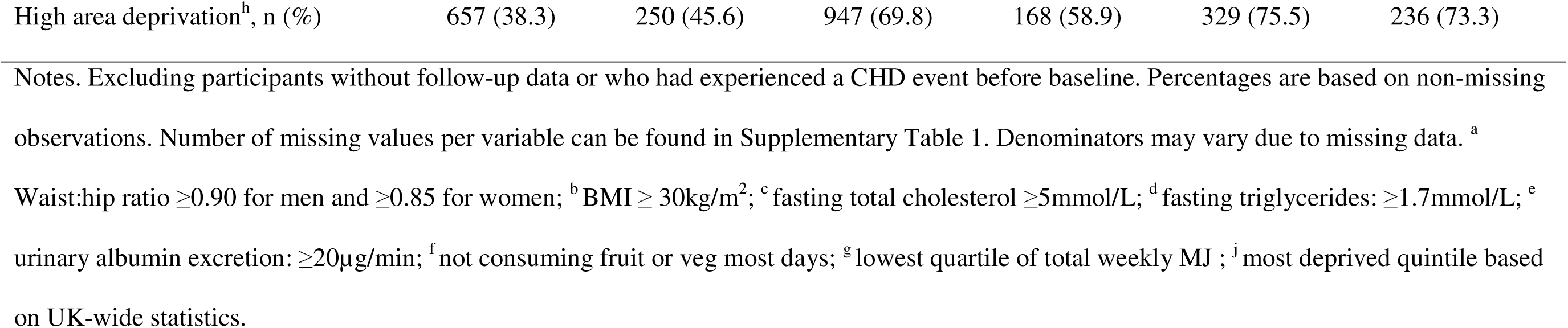
Analytical sample description at baseline.

|  | European |  | South Asian |  | African/African Caribbean |  |
| --- | --- | --- | --- | --- | --- | --- |
|  | Males | Females | Males | Females | Males | Females |
| N | 1751 | 551 | 1377 | 286 | 439 | 324 |
| Age (yrs), mean (SD) | 52.8 (7.1) | 52.9 (6.7) | 51.2 (7.0) | 50.7 (6.8) | 53.7 (5.8) | 53.0 (6.0) |
| Diabetes, n (%) | 126 (7.2) | 25 (4.5) | 306 (22.2) | 47 (16.4) | 79 (18.0) | 68 (21.0) |
| Hypertension, n (%) | 399 (22.8) | 114 (20.7) | 462 (33.6) | 78 (27.3) | 170 (38.8) | 157 (48.5) |
| Central adiposity<sup>a</sup>, n (%) | 1223 (69.8) | 111 (20.1) | 1222 (88.8) | 127 (44.6) | 322 (73.5) | 145 (44.8) |
| Obesity<sup>b</sup>, n (%) | 245 (14.0) | 92 (16.7) | 136 (9.9) | 74 (25.9) | 51 (11.6) | 120 (37.0) |
| Hypercholesterolemia<sup>c</sup>, n (%) | 1455 (83.2) | 442 (81.1) | 1128 (82.2) | 214 (75.6) | 304 (70.9) | 224 (71.1) |
| Hypertriglyceridemia<sup>d</sup>, n (%) | 689 (39.5) | 137 (25.2) | 739 (53.9) | 98 (34.8) | 88 (20.5) | 42 (13.3) |
| Albuminuria<sup>e</sup>, n (%) | 66 (6.4) | 12 (2.7) | 45 (6.1) | 7 (3.5) | 19 (6.8) | 23 (8.9) |
| Ever-smoker, n (%) | 1291 (73.7) | 287 (52.1) | 359 (26.1) | 7 (2.5) | 202 (46.3) | 48 (14.9) |
| Low fruit & veg consumption<sup>f</sup>, n (%) | 559 (31.9) | 148 (27.0) | 442 (32.3) | 64 (22.6) | 171 (39.5) | 63 (19.6) |
| Low physical activity<sup>g</sup>, n (%) | 308 (17.6) | 185 (33.6) | 356 (25.9) | 154 (53.8) | 93 (21.2) | 81 (25.0) |
| High area deprivation<sup>h</sup>, n (%) | 657 (38.3) | 250 (45.6) | 947 (69.8) | 168 (58.9) | 329 (75.5) | 236 (73.3) |

Notes. Excluding participants without follow-up data or who had experienced a CHD event before baseline. Percentages are based on non-missing observations. Number of missing values per variable can be found in Supplementary Table 1. Denominators may vary due to missing data. <sup>a</sup>
Waist:hip ratio $\geq 0.90$ for men and $\geq 0.85$ for women; <sup>b</sup> BMI $\geq 30\text{kg/m}^2$ ; <sup>c</sup> fasting total cholesterol $\geq 5\text{mmol/L}$ ; <sup>d</sup> fasting triglycerides: $\geq 1.7\text{mmol/L}$ ; <sup>e</sup> urinary albumin excretion: $\geq 20\mu\text{g/min}$ ; <sup>f</sup> not consuming fruit or veg most days; <sup>g</sup> lowest quartile of total weekly MJ ; <sup>j</sup> most deprived quintile based on UK-wide statistics.

Marked heterogeneity in the prevalence of risk factors was observed across sex and ethnic groups (*Table 1*). Both South Asians and African/African Caribbeans had greater risks of diabetes, hypertension and obesity, and were more likely to reside in the most deprived quintile of neighbourhoods. The prevalence of obesity in the South Asian and Africans/African Caribbean groups was even higher when using ethnicity and sex-specific cutoffs related to diabetes risk (*Supplementary Table 2*). The male excess prevalence of diabetes and hypertension observed in Europeans was maintained in South Asians but reversed in African/African Caribbeans. Prevalence of hypercholesterolemia was lower in African/African Caribbeans compared to other groups, while prevalence of hypertriglyceridemia was elevated in South Asian groups compared to Europeans, and lower in African/African Caribbeans. Both ethnic minority groups were less likely to be ever smokers, markedly so for women. Compared to European females, South Asian females were more, and African/African Caribbean females less likely to be physically active. Low fruit and vegetable consumption was slightly less common in South Asian and African/African Caribbean females. Area deprivation was markedly more common in South Asian and Africans/African Caribbean groups.

### Comparing incidence of CHD across sex and ethnicity

Over a median follow-up time of 40.8 years (estimated using the reverse Kaplan-Meier method), 1,710 first CHD events and 670 MI first events were experienced. Cumulative incidence of CHD (*Figure 1*) was greatest in South Asian males (65% by age 90), followed by South Asian females (55%). CHD incidence in European males (52%) was marginally lower than South Asian females. European females, and both African/African Caribbean groups had similarly low cumulative incidence of CHD (∼24-31%). Cumulative incidence of first MI events (*Supplementary Figure 2A*) showed similar relative group differences: greatest in South Asian males (29% by age 90) followed by South Asian females (21%) and European males (19%). European females, and both African/African Caribbean groups had similarly low cumulative incidence of MI (∼5-11%).

**Figure 1.**
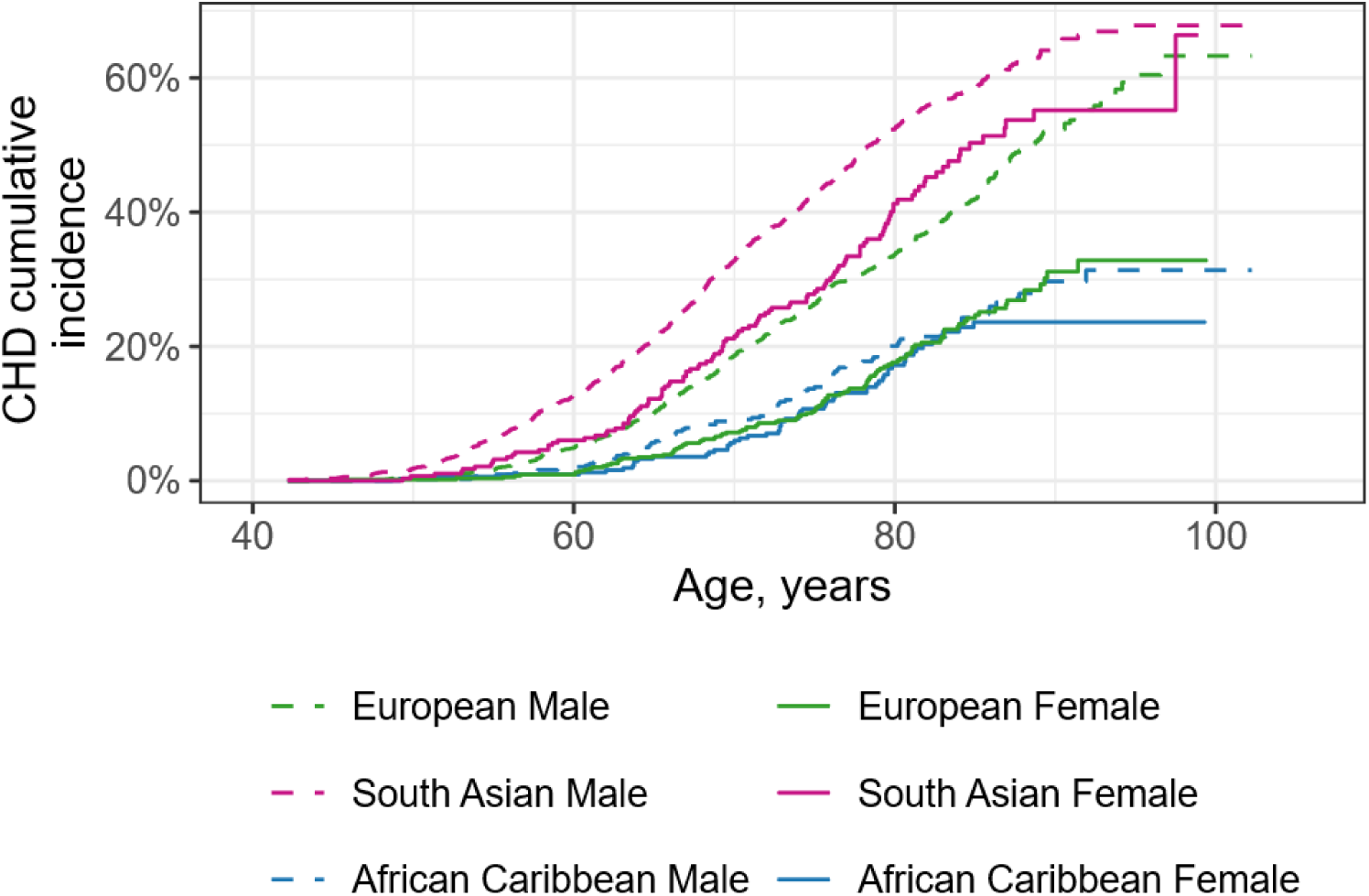
Cumulative incidence plots for coronary heart disease events by age. Coronary heart disease events represent both fatal and non-fatal events. Ethnicity indicated by line colour and sex indicated by line pattern. Number of coronary heart disease events/censored cases per group: European men (621/1,130), European women (111/440), South Asian men (707/670), South Asian women (119/167), African/African Caribbean men (95/344), African/African Caribbean women (57/267).

Over the follow-up period, 698 deaths due to CHD occurred. Cumulative incidence of CHD deaths (*Supplementary Figure 2B*) showed slightly different relative group differences: greatest in South Asian males (31% by age 90) followed by European males (24%). European females, South Asian females, and both African/African Caribbean groups had similarly low cumulative incidence of MI (∼6-14%).

Both sex and ethnicity were independently associated with risk of CHD (*HR [95% CI]: female v male: 0.51 [0.45, 0.58]; AFC v EUR: 0.52 [0.44, 0.62]; SA v EUR: 1.80 [1.63, 1.99]*), inclusion of the interaction term significantly improved model fit compared with a model containing only main effects (*likelihood ratio test; LRT: χ^2^(2) = 11.9, p = 0.003*). Sex and ethnicity were also independently associated with risk of MI (*HR [95% CI]: female v male: 0.43 [0.34, 0.54]; AFC v EUR: 0.34 [0.24, 0.48]; SA v EUR: 1.90 [1.62, 2.22]*), however inclusion of the interaction term did not significantly improve model fit compared with a model containing only main effects (*LRT: χ^2^(2) = 1.5, p = 0.472*). Similar patterns were observed for CHD death with significant main effects of sex and ethnicity (*HR [95% CI]: female v male: 0.43 [0.35, 0.54]; AFC v EUR: 0.37 [0.27, 0.50]; SA v EUR: 1.50 [1.28, 1.75]*), but inclusion of the interaction term did not significantly improve model fit (*LRT: χ^2^(2) = 1.0, p = 0.615*).

Sex differences in CHD differed by ethnicity. While they were clear in Europeans (*female v male: HR=0.45, 95% CI [0.37, 0.55]; Figure 2A*) they were markedly diminished in both South Asians (*HR=0.68, 95% CI [0.56, 0.82]*) and African/African Caribbeans (*HR=0.79, 95% CI [0.57, 1.10]*). However, the overall South Asian estimate masked a substantial time-varying effect of sex on CHD risk. In the first ten years of follow-up, South Asian females had markedly lower CHD hazard than males (*HR=0.35, 95% CI [0.21, 0.57]*), with this protective effect attenuating and reversing progressively over subsequent follow-up (10-20 years: *HR=0.77, 95% CI [0.56, 1.06]*; 20+ years: *HR=0.85, 95% CI [0.64, 1.14]*). No such time-varying pattern was identified in European or African/African Caribbean groups, for whom the proportional hazards assumption was satisfied, and a single estimate is reported.

**Figure 2.**
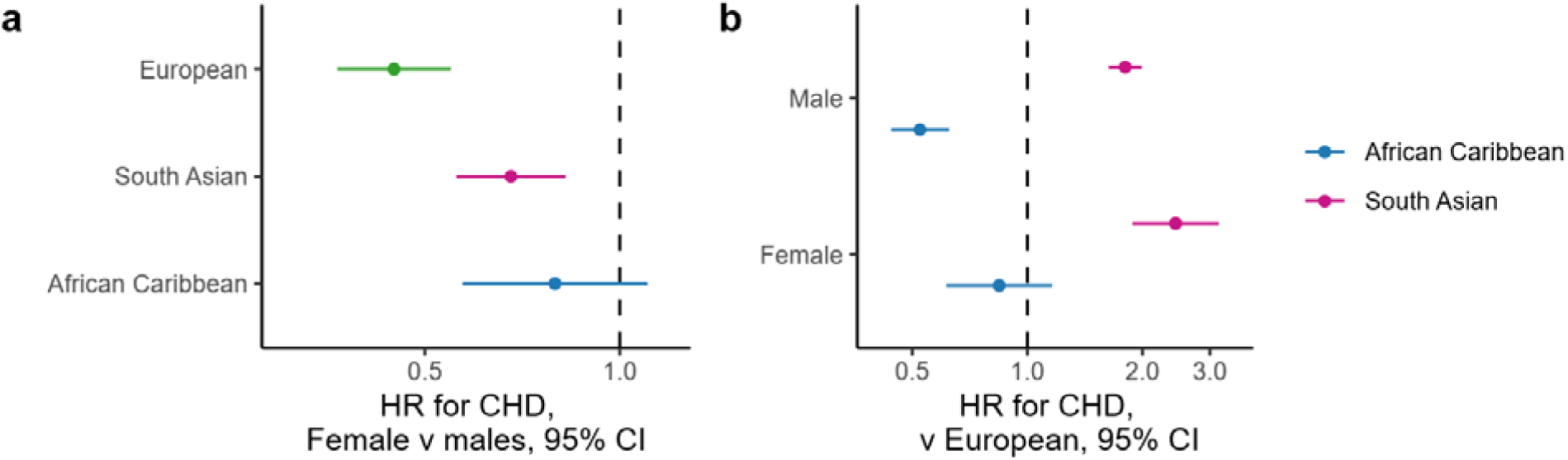
Hazard ratios (HRs) for coronary heart disease events by sex and ethnicity. HRs for ethnicity on coronary heart disease events stratified by sex. Comparisons are made against the European group. b) HRs for sex on CHD events stratified by ethnicity. HR derived from Cox proportional hazard models. All analyses control for age at baseline. N= 1,751 European men, 551 European women, 1,377 South Asian men, 286 South Asian women, 439 African/African Caribbean men, and 324 African/African Caribbean women.

Ethnic differences in CHD also differed by sex. In males, CHD risk was higher in South Asians compared to White Europeans (*HR=1.80, 95% CI [1.63, 1.99]*), and lower in African/African Caribbeans (*HR=0.52, 95% CI [0.44, 0.62]*). In females, the South Asian HR was even greater (*HR=2.44, 95% CI [1.88, 3.17]*), while African/African Caribbeans had no convincingly lower HR, although the estimate was imprecise (*HR=0.84, 95% CI [0.61, 1.16]*). Results were similar when modelling group differences using Fine-Grey models (Supplementary figure 3). Group differences in CHD deaths and in particular MI events were much smaller (Supplementary Figure 4).

### Risk factors for CHD across sex and ethnicity groups

The magnitude of the association between many risk factors and incident CHD was generally greater for females than males, most prominently for diabetes, elevated cholesterol, and elevated triglycerides (*Figure 3, Supplementary Table 3*). Magnitudes were especially high for South Asian females, with HRs for diabetes (*HR=2.38, 95% CI: 1.53,3.70*), elevated cholesterol and elevated triglycerides (*HR=2.06, 95% CI [1.27,3.35] and HR=1.81, 95% CI [1.25,2.63]* respectively); central adiposity (*HR=1.91, 95% CI: 1.31,2.78*), and hypertension (*HR=1.58, 95% CI [1.05,2.38]*) being the highest of any group.

**Figure 3.**
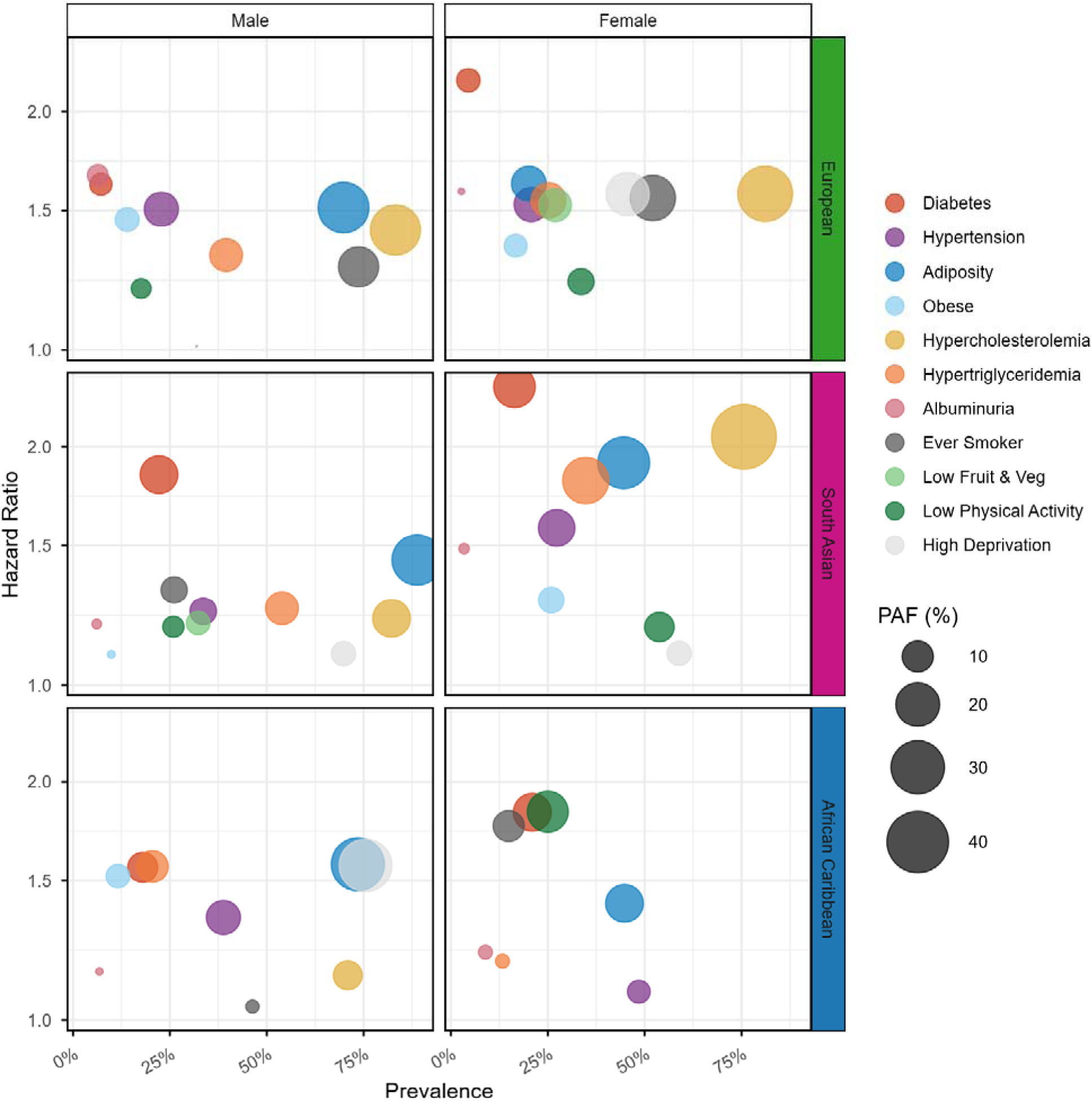
Prevalence, hazard ratios (HRs) and population attributable fractions (PAFs) for coronary heart disease plotted for risk factors by sex and ethnicity. Bubble size represents the PAF or relative contribution of each risk factor to coronary heart disease incidence per group, representing a combination of its group-wise risk and prevalence. Bubble colour represents the risk factor. Datapoints are not plotted where either the PAF fell below 0% or the HR fell below 1. HRs and PAFs derived from Cox proportional hazard models. PAFs calculated for a follow-up time of 20 years. All analyses control for age at baseline. N=1,751 European men, 551 European women, 1,377 South Asian men, 286 South Asian women, 439 African/African Caribbean men, and 324 African/African Caribbean women.

Some evidence of non-proportional hazards was observed (*Supplementary Table 3)*. Time-stratified analyses suggested that many were small differences and did not meaningful impact the patterns observed. Those with larger time-dependent differences were primarily driven by extreme follow-up times, where event counts were low and estimates were imprecise. The exception was for hypertriglyceridemia in European females (*pooled HR=1.55*). Time-stratified analyses showed a stronger association in the first 10 years (*HR=3.23*), attenuating in years 10-20 (*HR=1.70*) and approaching the null thereafter, although estimates at later follow-up were imprecise with wide confidence intervals.

In European males and females, the greatest population attributable fractions for CHD could be attributed to high prevalence, moderate-risk risk factors such as hypercholesterolemia (male: *PAF=26.4%, 95% CI [9.9, 42.9];* female: *31.7%, 95% CI [-2.6, 66.0]*) and ever smoking (male: *16.8%, 95% CI [4.9, 28.9];* female: *21.5%, 95% CI [3.2, 39.9]*) in addition to central adiposity in European males (*27.2%, 95% CI [16.2, 38.2])* and area deprivation in European females (1*9.6%, 95% CI [3.0, 36.2]; Figure 3, Supplementary Table 4*). These associations were similar upon further adjustment for confounders (*Supplementary Table 5)*.

In South Asian populations, metabolic risk factors such as diabetes, central adiposity and dyslipidaemia, accounted for more of the incident CHD risk. The impact of diabetes and of dyslipidaemia in South Asian females was especially strong compared to both South Asian and European men (diabetes: *18.0%, 95% CI [6.2, 29.8]* versus *14.8%, 95% CI [10.0, 19.7]* and *5.1%, 95% CI [1.6, 8.6]*; hypercholesterolemia: *44.2%, 95% CI [20.4, 68.1]* versus *14.7%, 95% CI [0.8, 28.7]* and *26.4%, 95% CI [9.9, 42.9]*; hypertriglyceridemia: *22.4%, 95% CI [7.9, 37.0]* versus *11.1%, 95% CI [3.6, 18.6]* and *11.1%, 95% CI [4.5, 17.7])*, and markedly higher than European females, reflecting the high risk these factors conferred. The same was true following adjustment for confounders (*Supplementary Table 5)*.

In African/African Caribbean groups, population attributable fractions were more modest. Although some exposures had elevated HRs (as noted above), their lower or more variable prevalence limited overall attributable burden. In African/African Caribbean men, the impact of central adiposity was on par with European and South Asian males (*29.7%, 95% CI [-1.5, 60.9]*) and the impact of area deprivation was particularly strong (*29.3%, 95% CI [-1.9, 60.5]*). In African/African Caribbean females there was also some suggestion that the association with low physical activity was greater than in other groups (*17.6%, 95% CI [0.1, 35.1]*). Interestingly, following adjustment for potential confounders, the contribution of central adiposity to CHD in African/African Caribbean females was also high (*27.1%, 95% CI [-2.9, 50.0]*). However, given the small number of CHD events in African/African Caribbean groups though, confidence limits around risk factor estimates were wide, and estimates themselves likely unstable.

## Discussion

We show that the well-established lower CHD incidence in females of European ancestry is largely absent for UK-resident females of South Asian and African/African Caribbean ancestry, when compared to males of the same ethnicity.^5^ Specifically, we find that while incident CHD HRs are halved in adult European females compared to European males, corresponding reductions for South Asian females are ∼a quarter, and African/African Caribbean females ∼a fifth compared to their male counterparts. Thus, South Asian females have close to a 2.5-fold excess CHD HR compared to European females, markedly larger than the excess South Asian versus European male HR at 1.6. These relationships manifest in a particularly high incidence of CHD in South Asian females, above even that experienced by White European males. These findings point towards important and frequently overlooked intersectional differences in CHD risk, and support the imperative for more granularity in such research.^4^

Interestingly, time-stratified analyses indicate that the reduced protection for the South Asian female group does not occur until older age (>10 years post-baseline, mean age>62 years). This may reflect a more rapid accumulation of risk factors in later life, worsening of existing health problems or greater sensitivity to age-specific events such as the menopause that confer greater risk for CHD.^31–33^ Longitudinal assessment of time-varying risk factors is needed to explore this further.

In comparison, sex-by-ethnicity differences in MI events or CHD deaths were not as clear and sex differences in South Asian and African/African Caribbean groups were more similar to those seen in Europeans. Research using European registries suggests that first-generation migrants from South Asian or African countries also experience smaller sex differences in MI incidence compared to locally-born populations matching the direction of effect in this study, however these benefited from larger populations in which to reliably detect small effects ^9,10^. While smaller numbers of events preclude any strong conclusions being drawn, these results potentially reflect stronger intersectional variation in non-fatal presentations of CHD, such as angina or stable disease, rather than acute or fatal coronary events.^18^ Unfortunately, minimal research has examined intersectional differences in sub-acute CHD outcomes, favouring isolated effects,^34^ however research indicates that sex differences in prognosis following acute coronary syndrome are greater in South Asian groups compared to Europeans, potentially supporting our findings.^35^

Ethnicity itself is not a modifiable determinant of CHD risk, rather it is associated with a plethora of behavioural, socioeconomic and biological factors which confer CVD risk.^4^ Ethnic and sex differences in both the distribution or impact of cardiovascular risk factors are key candidates to account for the loss of CHD risk protection in ethnic minority females. South Asian females exhibit a disproportionately high burden of CVD risk factors with higher prevalence of diabetes, hypertension and central adiposity than white European males, accompanied by similar levels of dyslipidaemia (both excess triglycerides and total cholesterol). These patterns, alongside the particularly high risk these factors confer on CHD outcomes are reflected in the PAFs which indicate the relative importance of a given risk factor in its association with disease. PAFs in South Asian females for dyslipidaemia, diabetes and central adiposity were amongst the highest estimated, reflecting both their high prevalence and greater risk impact compared to the other groups. This risk profile potentially overwhelms the impact of smoking, given its relatively low prevalence in this group, a pattern mirrored in US populations.^32^ Crucially, our work shows that these risk factors appear to confer a greater relative risk for CHD in South Asian females than other subgroups, an observation previously seen in South Asians men and women combined,^3^ but here extended to sex-specific groups.

Despite their low risk of CHD, African/African Caribbean females have one of the highest prevalences of diabetes, hypertension, and albuminuria, reversing the sex difference in risk factor prevalence generally observed in white Europeans. African/African Caribbean females were the most obese of all six subgroups, with a prevalence over three times that of African/African Caribbean males. This combination of prevalent risk factors in women likely determines the abrogation of CHD protection that is commonly observed in African/African Caribbean males.

### Strengths and limitations

This study has several strengths. It benefits from a comprehensive assessment of both baseline risk factors and incident CHD events over a long follow-up time of over 20 years. Risk factors were directly assessed, allowing for a more comprehensive assessment of metabolic, cardiovascular and environmental health, than could be ascertained through linked records. We were able to accurately ascertain CHD events through a combination of linked records and clinician assessment, avoiding the potential misreporting of self-reported health as well as minimisation of participant attrition which often biases research into sex and ethnicity.^36^ The SABRE cohort also benefits from a high response rate (63%),^17^ resulting in substantially greater representativeness than larger volunteer cohorts with very low participation and limited minority ethnic inclusion.^37^ This reduces the risk of participation bias and strengthens the validity of analyses of ethnic inequalities in risk, and in exposure-outcome relationships.

Some limitations should be noted. Intersectional approaches to health research frequently observe poorer health outcomes for ethnic minority women, emphasising the importance of considering multiple characteristics simultaneously. However, it should be acknowledged that this study only considered two grouping factors (ethnicity and sex) and other factors, such as socioeconomic status ^38^ and comorbidity may interact to lead to further inequalities in CHD outcomes.^39^ Future research, with larger cohorts, should aim to examine these intersectional states. Some sex-ethnicity groups were relatively small, particularly the African and African Caribbean women, leading to wide confidence intervals and limiting the analyses that could be performed and conclusions that could be made. Due to the original aims of the cohort’s study design, we were limited to first generation migrant ethnic minority groups who may experience significantly different health profiles than subsequent generations due to differences in early and later life exposomes.^40^ We were also unable to explore the cardiovascular outcomes of finer grained ethnic subgroups. The South Asian groups were wholly of Indian origin, and we had to combine those who migrated from African and Caribbean countries, ignoring the fact that ancestral subgroups have different distributions of risk factors and risks of disease.^41^ Finally, we were only able to examine risk factors present at baseline. Despite the provision of universal free healthcare in UK, ethnic minorities frequently experience poorer healthcare and may have experienced poorer management of early cardiovascular signs such as hypertension.

### Conclusion

Ethnic minority women of South Asian or African/African Caribbean ancestry in the UK do not have the same protection from CHD as their white European counterparts. Indeed, South Asian females have CHD risks that are greater than white European males, despite negligible smoking exposure. In females, we find important between ethnic group differences in risk factor distribution, and in the association of risk factors with outcomes, suggesting a different CHD phenotype to that observed in Europeans. We have previously reported poor predictive ability of CVD risk scores, designed to determine which individuals should get preventative medication, and show that ethnic minority women run the risk of being both under and overtreated.^43^ Our findings are a call for greater representation of women from different ethnic groups in population cohorts and examination of explanations for differences in sex distribution of risk factors, their associations with ethnicity and with outcomes.

## Acknowledgments

We are extremely grateful to all the people who took part in the study, and past and present members of the SABRE team who helped to collect and analyse the data. This work uses data provided by patients and collected by the NHS as part of their care and support. NHS England (formerly NHS Digital) provided linked Hospital Episode Statistics (HES) and mortality records under the Data Sharing Framework Contract and associated Data Sharing Agreement. NHS England bears no responsibility for the analysis, interpretation or conclusions contained in this publication. The data were accessed securely in accordance with the Data Sharing Framework Contract and SABRE governance procedures.

## Data availability statement

SABRE data are available to bona fide researchers upon request. Data sharing is managed by the study team at UCL, and access requires submission of a data-sharing application and approval under the study’s governance procedures.

