## Supplemental Information for "Sex and ethnicity differences in coronary heart disease: A UK-based tri-ethnic cohort analysis"

### Supplementary Methods

#### Defining coronary heart disease events

Coronary heart disease (CHD) events were identified from ONS mortality records and NHS hospital episode statistics using ICD-9 codes: 410, 411, 412, 413 and 414; and ICD-10 codes: I20, I21, I22, I23, I24, I25, Z951, and Z955. The event date was taken as the date of mortality or the date of admission to hospital. Primary care data were reviewed independently by 2 senior physicians blinded to participant identity or characteristics. A CHD event was defined if both physicians agreed on definite or probable diagnosis of myocardial infarction or acute coronary syndrome, according to pre-determined criteria used in the ASCOT (Anglo-Scandinavian Cardiac Outcomes Trial).^1^ This was based on recorded symptoms, cardiac enzymes, electrocardiography findings, and hospital discharge diagnosis. Adjudication by a third physician was conducted if required.

CHD-related deaths were identified solely from ONS mortality records using ICD-9 codes: 410, 411, 412, 413 and 414; and ICD-10 codes: I20, I21, I22, I23, I24, and I25.

Myocardial infarction (MI) events were identified from ONS mortality records and NHS hospital episode statistics using ICD-9 codes: 410, and ICD-10 codes: I21 and I22. The event date was taken as the date of mortality or the date of admission to hospital. Primary care data were reviewed as described above. An MI event was defined if both physicians agreed on definite or probable diagnosis of myocardial infarction.

#### Ethnicity-specific cutoffs

Ethnicity-specific measures of obesity and adiposity were also investigated in relation to sex and ethnic differences in CHD. Ethnicity-specific central adiposity was defined using ethnicity specific waist circumference cut-offs as defined previously (European men: 102 cm, South Asian men: 90.4 cm, African/African Caribbean men: 90.6 cm, European women: 88 cm, South Asian women: 84.0 cm, African/African Caribbean women: 81.2 cm).^2^ Ethnicity-specific obesity was also defined using ethnicity specific cut-offs as defined previously (European: >30kg/m^2^, South Asia: >25.2kg/m^2^, African/African Caribbean: >27.2kg/m^2^).^2^ Variables were used in the same analyses as the universal obesity and adiposity cutoffs (Supplementary Table 2).

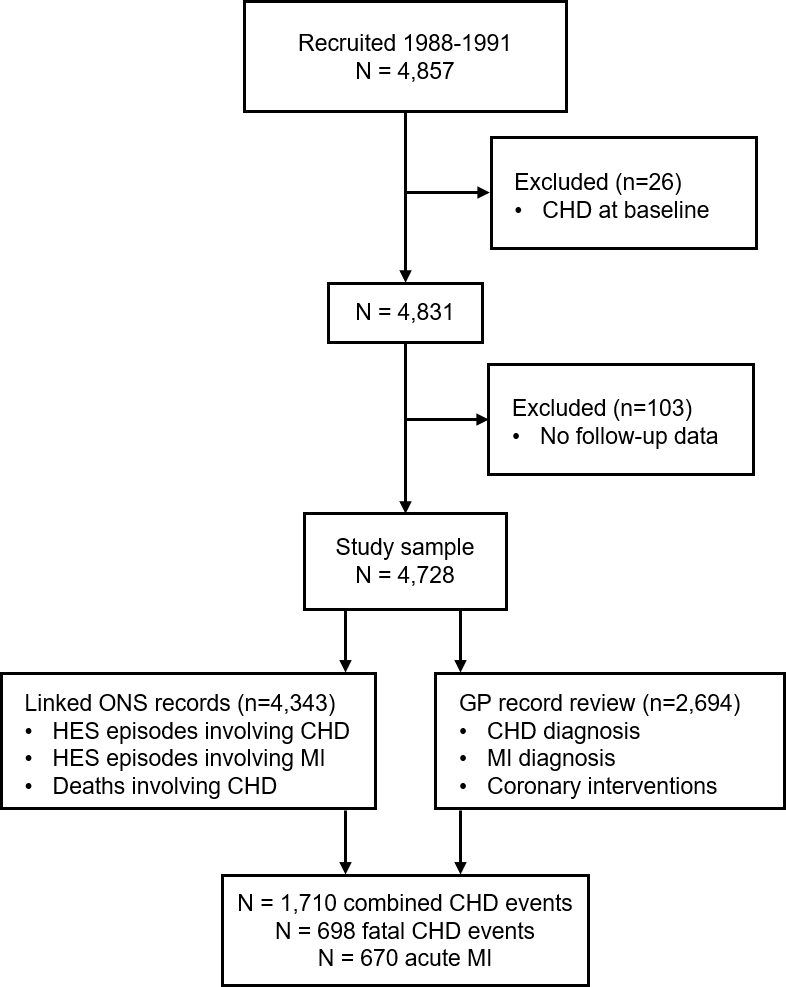

### Supplementary Figures

**Supplementary Figure 1. Cohort flowchar****t**

CHD: coronary heart disease, MI: myocardial infarction, ONS: Office for National Statistics, GP: general practitioner.

**Supplementary Figure 2. Cumulative incidence plots for myocardial infarction (MI) events and coronary heart disease (CHD) deaths by age.**

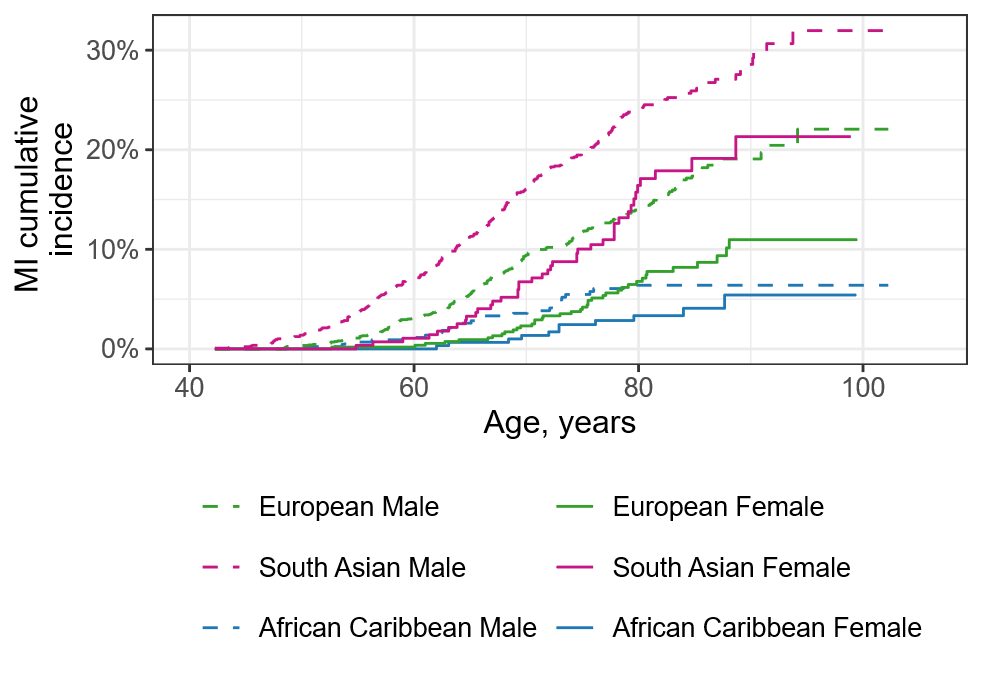

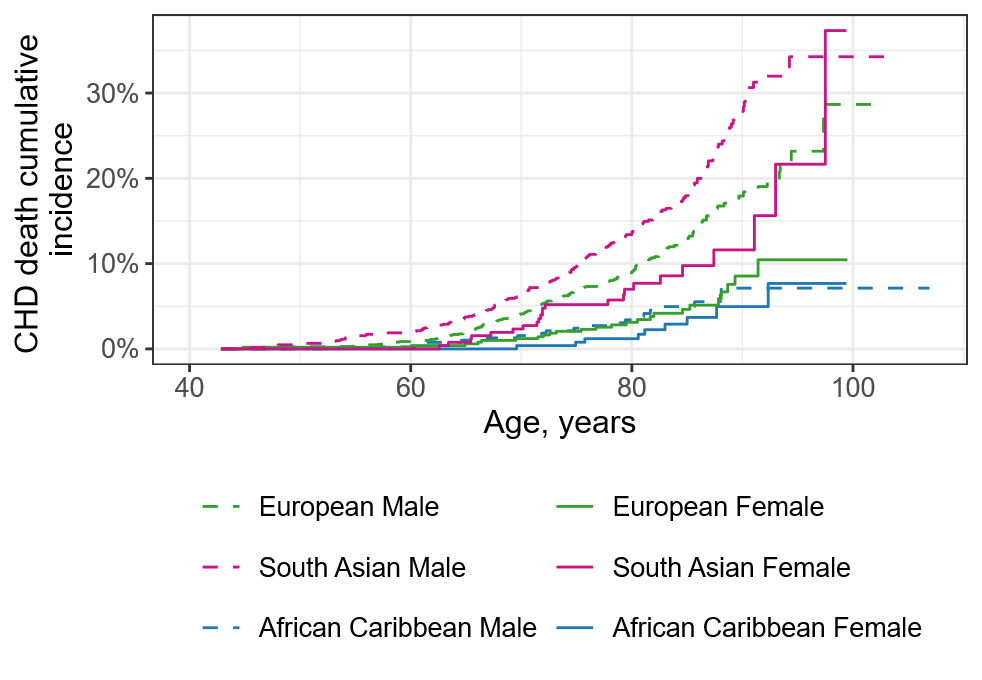

**a**

b

MI events represent both fatal and non-fatal events. Ethnicity indicated by line colour and sex indicated by line pattern.

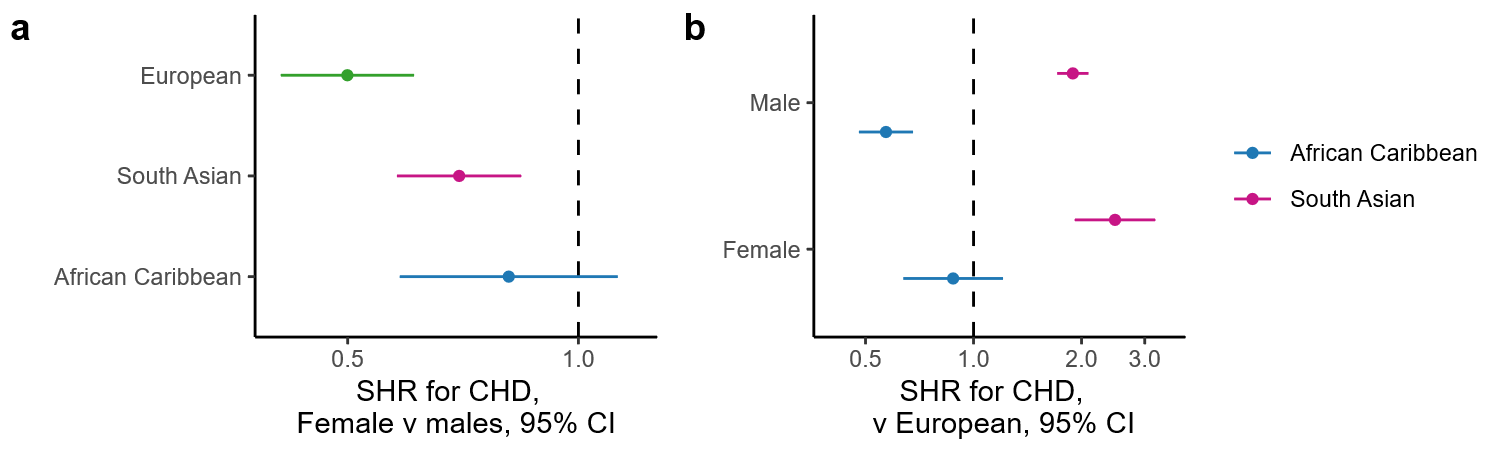

**Supplementary Figure 3. Supplementary Figure 3. Sub-hazard ratios (SHRs) for coronary heart disease (CHD) events by sex and ethnicity.**

SHRs derived using Fine-Grey models using non-CHD-related death as a competing event. a) SHRs for ethnicity on CHD events stratified by sex. Comparisons are made against the European group. b) SHRs for sex on CHD events stratified by ethnicity. All analyses control for age at baseline.

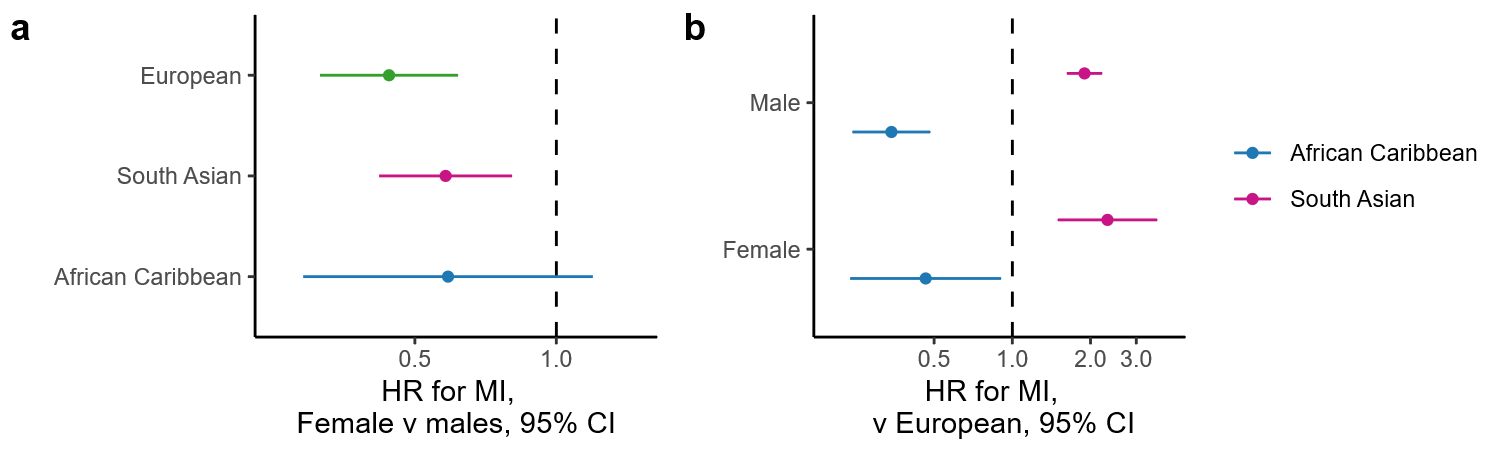

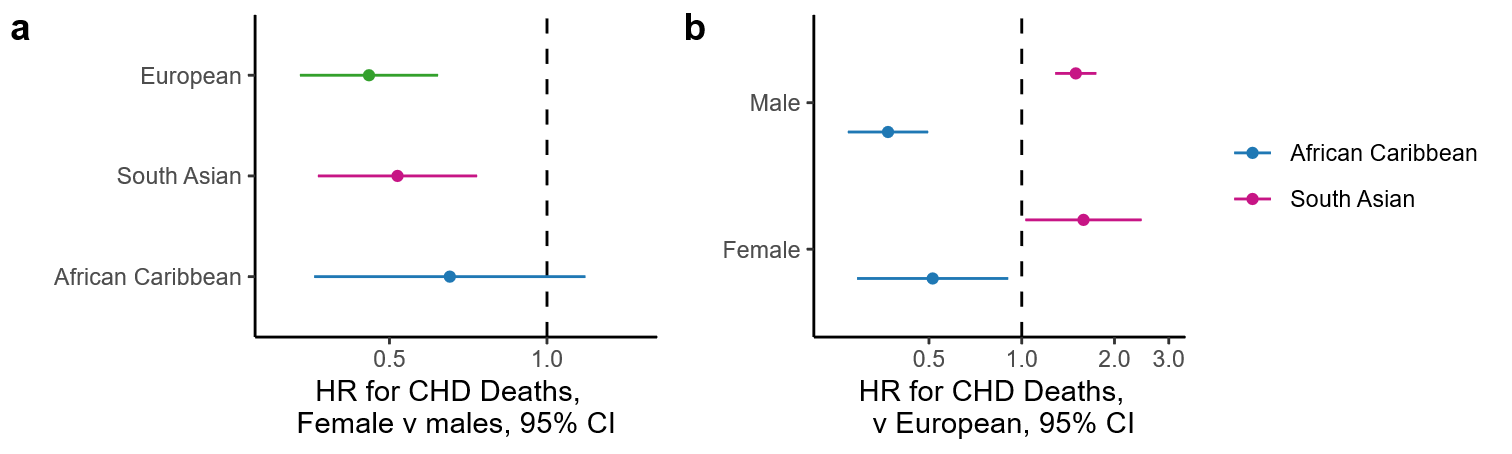

d

**c**

**Supplementary Figure 4. Hazard ratios (HRs) for myocardial infarction (MI) events and coronary heart disease (CHD) deaths by sex and ethnicity.**

a) HRs for ethnicity on MI events stratified by sex. Comparisons are made against the European group. b) HRs for sex on MI events stratified by ethnicity. c) HRs for ethnicity on CHD deaths stratified by sex. Comparisons are made against the European group. d) HRs for sex on CHD deaths stratified by ethnicity. All analyses control for age at baseline.

### Supplementary Tables

| Supplementary Table 1. Missing values in baseline characteristics, n (%) | | | | | | |
| --- | --- | --- | --- | --- | --- | --- |
|  | **European** | | **South Asian** | | **African Caribbean** | |
|  | **Males** | **Females** | **Males** | **Females** | **Males** | **Females** |
| N | 1752 | 551 | 1377 | 286 | 439 | 324 |
| Age | 0 (0) | 0 (0) | 0 (0) | 0 (0) | 0 (0) | 0 (0) |
| Diabetes | 0 (0) | 0 (0) | 0 (0) | 0 (0) | 0 (0) | 0 (0) |
| Hypertension | 2 (0.1) | 0 (0) | 0 (0) | 0 (0) | 1 (0.2) | 0 (0) |
| Central adiposity | 1 (0.1) | 0 (0) | 1 (0) | 1 (0) | 1 (0.2) | 0 (0) |
| Obesity | 2 (0.1) | 0 (0) | 2 (0.1) | 0 (0) | 1 (0.2) | 0 (0) |
| Hypercholesterolemia | 4 (0.2) | 6 (1.1) | 5 (0.4) | 3 (1.0) | 10 (2.3) | 9 (2.8) |
| Hypertriglyceridemia | 5 (0.3) | 7 (1.3) | 6 (0.4) | 4 (1.4) | 10 (2.3) | 9 (2.8) |
| Albuminuria | 721 (41.2) | 113 (20.5) | 636 (46.2) | 83 (29.0) | 161 (36.7) | 66 (20.4) |
| Ever smoker | 1 (0.1) | 0 (0) | 3 (0.2) | 2 (0.7) | 3 (0.7) | 2 (0.6) |
| Low fruit & veg consumption | 0 (0) | 1 (0.2) | 9 (0.7) | 3 (1.0) | 6 (1.4) | 3 (0.9) |
| Low physical activity | 0 (0) | 0 (0) | 0 (0) | 0 (0) | 0 (0) | 0 (0) |
| High area deprivation | 37 (2.1) | 3 (0.5) | 20 (1.5) | 1 (0.3) | 3 (0.7) | 2 (0.6) |

| Supplementary Table 2. Results for ethnic group-specific categorisation of central adiposity and obesity | | | | | | |
| --- | --- | --- | --- | --- | --- | --- |
|  | European | | South Asian | | African Caribbean | |
|  | Males | Females | Males | Females | Males | Females |
| Description, n (%) |  |  |  |  |  |  |
| Group-specific central adiposity | 268 (15.3) | 102 (18.5) | 804 (58.4) | 131 (46.0) | 192 (43.8) | 221 (68.2) |
| Group-specific obesity | 245 (14.0) | 92 (16.7) | 770 (56.0) | 178 (62.2) | 172 (39.3) | 227 (70.1) |
| Association with CHD (HR [95% CI]) |  |  |  |  |  |  |
| Group-specific central adiposity | 1.47 [1.20,1.79] | 1.60 [1.05,2.46] | 1.24 [1.06,1.45] | 1.29 [0.89,1.86] | 1.33 [0.89,1.99] | 1.62 [0.87,3.02] |
| Group-specific obesity | 1.46 [1.19,1.80] | 1.35 [0.86,2.14] | 1.21 [1.05,1.41] | 1.46 [0.99,2.15] | 1.42 [0.95,2.13] | 1.23 [0.67,2.25] |
| Population attributable fraction for CHD (% [95% CI]) |  |  |  |  |  |  |
| Group-specific central adiposity | 7.2 [3.0,11.4] | 10.4 [0.1,20.8] | 12.1 [4.1,20.1] | 11.6 [-5.0,28.2] | 12.6 [-5.5,30.6] | 30.1 [-3.5,30.6] |
| Group-specific obesity | 5.9 [1.9,9.8] | 5.6 [-3.8,14.9] | 10.3 [2.5,18.1] | 22.0 [-0.2,44.2] | 14.0 [-2.1,30.2] | 14.3 [-23.7,52.2] |
| Notes. HR=hazard ratio, CI=confidence interval. | | | | | | |

| Supplementary Table 3. Hazard ratios [95% CI] for the impact of CHD risk factors on CHD events within sex and ethnicity sub-groups. | | | | | | |
| --- | --- | --- | --- | --- | --- | --- |
|  | European | | South Asian | | African/African Caribbean | |
|  | Males | Females | Males | Females | Males | Females |
| Diabetes | 1.62 [1.24,2.11] | 2.19 [1.10,4.35] | 1.84 [1.56,2.19] | 2.38 [1.53,3.70] | 1.56 [0.96,2.54] | 1.83 [1.02,3.27] |
| Hypertension | 1.51 [1.26,1.80] | 1.53 [1.00,2.33] | 1.24 [1.06,1.45] | 1.58 [1.06,2.38] | 1.35 [0.90,2.02] | 1.09 [0.64,1.84] |
| Central adiposity | 1.51 [1.25,1.83] | 1.62 [1.07,2.45] | 1.44 [1.12,1.87] | 1.91 [1.31,2.78] | 1.57 [0.94,2.64] | 1.40 [0.83,2.38] |
| Obesity | 1.46 [1.19,1.80] | 1.35 [0.86,2.14] | 1.09 [0.86,1.39] | 1.28 [0.86,1.92] | 1.52 [0.89,2.61] | 0.73 [0.42,1.27] |
| Hypercholesterolemia | 1.42 [1.12,1.80] | 1.58 [0.85,2.93] | 1.21 [0.99,1.48] | 2.06 [1.27,3.35] | 1.14 [0.73,1.86] | 0.77 [0.44,1.37] |
| Hypertriglyceridemia | 1.32 [1.13,1.54] | 1.55 [1.04,2.31] | 1.25 [1.08,1.45] | 1.81 [1.25,2.63] | 1.56 [0.98,2.50] | 1.19 [0.56,2.51] |
| Albuminuria | 1.66 [1.17,2.37] | 1.59 [0.50,5.04] | 1.20 [0.79,1.80] | 1.49 [0.53,4.16] | 1.15 [0.41,3.27] | 1.22 [0.43,3.43] |
| Ever-smoker | 1.27 [1.06,1.53] | 1.56 [1.07,2.27] | 1.32 [1.12,1.55] | 0.22 [0.03,1.57] | 1.04 [0.69,1.56] | 1.76 [0.91,3.40] |
| Low fruit & veg consumption | 1.01 [0.85,1.20] | 1.52 [1.02,2.29] | 1.20 [1.02,1.40] | 0.81 [0.52,1.27] | 0.78 [0.51,1.19] | 0.71 [0.34,1.51] |
| Low physical activity | 1.20 [0.98,1.46] | 1.22 [0.83,1.79] | 1.19 [1.00,1.41] | 1.18 [0.82,1.72] | 0.98 [0.59,1.63] | 1.83 [1.06,3.17] |
| High area deprivation | 0.98 [0.83,1.15] | 1.57 [1.08,2.30] | 1.10 [0.93,1.30] | 1.10 [0.76,1.58] | 1.57 [0.91,2.70] | 0.83 [0.47,1.47] |
| Note. All models adjusted for age at baseline. | | | | | | |

| Supplementary Table 4. Population attributable fractions (%, [95% CI]) for the impact of CHD risk factors on CHD events within sex and ethnicity sub-groups. | | | | | | |
| --- | --- | --- | --- | --- | --- | --- |
|  | European | | South Asian | | African/African Caribbean | |
|  | Males | Females | Males | Females | Males | Females |
| Diabetes | 5.1 [1.6,8.6] | 5.6 [-3.5,14.7] | 14.8 [10.0,19.7] | 18.0 [6.2,29.8] | 9.3 [-1.5,20.1] | 14.9 [-1.9,31.7] |
| Hypertension | 12.0 [6.2,17.7] | 12.0 [-1.7,25.7] | 7.2 [1.7,12.8] | 13.9 [1.6,26.2] | 12.0 [-5.0,29.1] | 5.2 [-21.4, 32.0] |
| Central adiposity | 27.2 [16.2,38.2] | 12.4 [0.5,24.4] | 26.6 [9.8,43.5] | 28.2 [12.0,44.5] | 29.7 [-1.5,60.9] | 14.7 [-9.5,38.9] |
| Obesity | 5.9 [2.4,9.5] | 5.4 [-4.0,14.7] | 0.8 [-1.6,3.3] | 6.7 [-4.3,17.8] | 5.8 [-2.7,14.2] | -11.4 [-30.1, 7.2] |
| Hypercholesterolemia | 26.4 [9.9,42.9] | 31.7 [-2.6,66.0] | 14.7 [0.8,28.7] | 44.2 [20.4,68.1] | 8.7 [-22.5,39.8] | -20.5 [-66.0,25.0] |
| Hypertriglyceridemia | 11.1 [4.5,17.7] | 12.7 [-0.9,26.4] | 11.1 [3.6,18.6] | 22.4 [7.9,37.0] | 10.3 [-1.3,22.0] | 2.1 [-9.4,13.5] |
| Albuminuria | 4.5 [0.6,8.3] | 0.7 [-4.9,6.3] | 1.1 [-2.2,4.5] | 1.3 [-2.4,4.9] | 0.8 [-9.8,11.3] | 2.1 [-8.4,12.5] |
| Ever-smoker | 16.8 [4.9,28.6] | 21.5 [3.2,39.9] | 7.1 [2.4,11.8] | -2.3 [-5.0,0.3] | 1.9 [-17.4,21.2] | 10.0 [-4.9,24.8] |
| Low fruit & veg consumption | 0.4 [-4.9,5.8] | 11.0 [-0.6,22.7] | 5.7 [0.8,10.6] | -4.6 [-13.2,4.1] | -9.3 [-24.8,6.1] | -6.1 [-18.2,6.0] |
| Low physical activity | 4.1 [-0.5,8.7] | 7.0 [-7.2,21.2] | 4.7 [-0.2,9.5] | 9.0 [-9.5,27.5] | -0.1 [-11.3,11.1] | 17.6 [0.1,35.1] |
| High area deprivation | -0.8 [-7.3,5.6] | 19.6 [3.0,36.2] | 6.1 [-4.6,16.9] | 6.1 [-15.6,27.7] | 29.3 [-1.9,60.5] | 2.5 [-59.2,31.1] |
| Note. All models adjusted for age at baseline. Calculated at follow-up time of 20 years. | | | | | | |

| Supplementary Table 5. Population attributable fractions (%, [95% CI]) for the impact of CHD risk factors on CHD events within sex and ethnicity sub-groups. | | | | | | |
| --- | --- | --- | --- | --- | --- | --- |
|  | European | | South Asian | | African/African Caribbean | |
|  | Males | Females | Males | Females | Males | Females |
| ***Adjusting for area deprivation*** | | | | | | |
| Diabetes | 5.3 [1.6,9.0] | 4.7 [-3.7,13.2] | 16.0 [10.7,21.2] | 19.0 [5.9,32.1] | 9.7 [-2.1,21.5] | 14.1 [-2.5,30.8] |
| Hypertension | 12.4 [6.7,18.2] | 12.6 [-1.4,26.5] | 7.6 [1.8,13.5] | 15.2 [0.7,28.9] | 11.0 [-5.6,26.2] | 5.1 [-23.5, 33.7] |
| Central adiposity | 28.0 [16.9,39.0] | 10.7 [-2.3,23.8] | 28.5 [10.8,46.2] | 28.9 [12.5,45.2] | 27.6 [-5.8,58.2] | 19.1 [-4.9,43.2] |
| Obesity | 6.2 [2.5,10.0] | 4.5 [-4.9,14.0] | 0.9 [-1.9,3.6] | 6.1 [-5.1,17.4] | 4.9 [-3.9,13.6] | -11.6 [-31.7, 8.5] |
| Hypercholesterolemia | 26.1 [10.7,41.6] | 36.0 [-2.9,75.0] | 16.4 [2.4,30.4] | 45.1 [21.5,68.7] | 7.0 [-26.5,40.5] | -18.2 [-67.8,31.4] |
| Hypertriglyceridemia | 10.9 [4.5,17.4] | 11.4 [-3.1,25.8] | 11.0 [3.4,18.6] | 21.9 [7.4,36.4] | 9.7 [-2.3,21.7] | 2.4 [-9.6,14.5] |
| Albuminuria | 4.2 [0.0,8.4] | 1.0 [-5.5,7.5] | 1.3 [-2.5,5.1] | 1.3 [-3.1,5.6] | 1.7 [-8.2,11.7] | 1.8 [-9.4,13.1] |
| Ever-smoker | 17.9 [5.3,30.6] | 20.8 [2.3,39.3] | 7.0 [2.2,11.8] | -1.9 [-4.1,0.4] | 1.6 [-19.0,22.1] | 11.6 [-4.4,27.5] |
| Low fruit & veg consumption | 0.9 [-4.4,6.2] | 10.0 [-2.3,22.2] | 5.2 [-0.3,10.8] | -5.0 [-13.7,3.7] | -9.8 [-26.3,6.6] | -5.8 [-18.7,7.1] |
| Low physical activity | 3.9 [-1.0,889] | 8.4 [-7.0,23.8] | 5.4 [0.0,10.8] | 7.7 [-13.9,29.3] | -1.5 [-12.4,9.4] | 19.8 [1.2,38.3] |
| ***Additionally adjusting for smoking, fruit & veg consumption and physical activity*** | | | | | | |
| Diabetes | 5.0 [1.1,8.8] | 3.7 [-5.2,12.6] | 15.7 [10.2,21.1] | 18.9 [6.2,32.1] | 9.2 [-3.3,21.6] | 15.5 [-1.8,32.8] |
| Hypertension | 12.9 [6.9,18.9] | 15.8 [1.2,30.4] | 8.3 [2.1,14.4] | 15.9 [0.7,31.1] | 10.3 [-6.7,27.2] | 4.0 [-25.4, 33.4] |
| Central adiposity | 26.6 [15.7,37.5] | 10.9 [-1.6,23.4] | 28.9 [11.4,46.5] | 31.5 [14.6,48.3] | 27.2 [-2.4,57.5] | 17.3 [-6.5,41.1] |
| Obesity | 5.9 [2.0,9.9] | 4.6 [-4.9,14.0] | 0.6 [-2.2,3.3] | 4.7 [-8.2,17.5] | 5.4 [-3.7,14.5] | -12.7 [-32.2, 6.9] |
| Hypercholesterolemia | 25.3 [9.3,41.4] | 34.9 [-4.9,74.7] | 17.0 [2.8,31.3] | 48.0 [26.1,69.9] | 6.1 [-24.5,36.7] | -18.5 [-68.7,31.8] |
| Hypertriglyceridemia | 9.7 [2.9,16.5] | 9.4 [-5.9,24.7] | 11.4 [3.6,19.2] | 22.6 [7.4,37.8] | 10.0 [-2.8,22.8] | 1.8 [-10.4,14.0] |
| Albuminuria | 4.3 [0.0,8.7] | 0.9 [-5.9,7.7] | 1.2 [-2.6,4.9] | 1.5 [-3.2,6.2] | 0.3 [-12.7,13.2] | 0.3 [-12.7,13.2] |
| ***Additionally adjusting for obesity*** | | | | | | |
| Diabetes | 4.8 [1.2,8.4] | 3.9 [-5.3,13.0] | 15.4 [9.9,20.9] | 18.9 [5.8,32.0] | 8.2 [-4.0,20.4] | 15.2 [-4.3,34.7] |
| Hypertension | 11.9 [5.7,18.2] | 14.0 [-0.7,28.7] | 7.8 [1.7,13.9] | 15.7 [0.7,30.6] | 9.2 [-7.9,26.4] | 6.3 [-24.9, 27.1] |
| Central adiposity | 24.0 [11.9,36.1] | 9.9 [-3.5,23.3] | 28.2 [10.8,45.6] | 31.0 [14.6,47.3] | 24.7 [-9.5,58.9] | 27.1 [-2.9,50.0] |
| Hypercholesterolemia | 25.2 [8.4,42.1] | 34.0 [-5.1,73.1] | 16.6 [2.0,31.2] | 43.8 [25.7,70.4] | 4.5 [-27.9,36.9] | -20.5 [-72.1,31.2] |
| Hypertriglyceridemia | 8.0 [0.4,15.6] | 9.3 [-5.2,23.7] | 11.1 [3.2,19.0] | 21.9 [6.6,37.2] | 8.1 [-5.3,21.5] | 3.7 [-8.6,16.0] |
| Albuminuria | 3.7 [-0.4,7.9] | 0.9 [-5.7,7.5] | 1.0 [-2.4,4.4] | 1.2 [-3.7,6.1] | 0.7 [-115,12.9] | -0.7 [-13.9,12.6] |
| Note. All models adjusted for age at baseline. | | | | | | |
